# Evaluating the accuracy and consistency of ChatGPT for the management of type 2 diabetes: A cross-sectional study

**DOI:** 10.1101/2025.07.04.25330845

**Authors:** Danielle Cutler, Tamara Van Bakel, Patricia Olar, Michael Fralick

**Author notes:** **Correspondence:** Michael Fralick, MD, PhD, SM, FRCPC, Sinai Health System, 60 Murray Street, Toronto, ON M5T 3L9. Authorship statement Conceptualization: All authors Data Curation: All authors Formal Analysis: Cutler Methodology: All authors Supervision: Fralick Writing original draft: Cutler & Fralick Writing review & Editing: All authors.

## Abstract

Large language models (LLMs) have fundamentally changed how patients and clinicians retrieve information; however, it is unclear how accurate and consistent widely available LLMs are in answering questions related to medical information. Our objective was to evaluate the accuracy and consistency of ChatGPT in answering questions related to the management of type 2 diabetes mellitus (T2DM). Three users asked ChatGPT 13 questions pertaining to medications from the top five most common classes of T2DM medications. A response was labelled inconsistent if the response provided to one user differed from the response provided to at least one other user in the same domain for the same medication. A response was labelled as inaccurate if the information provided by ChatGPT was incorrect based on the most recent FDA-approved drug label, in addition to review by an expert reviewer. Additionally, one user asked ChatGPT 26 basic questions related to the management of T2DM, in which the answer was categorized as correct or incorrect. We summarized all results using descriptive statistics. ChatGPT delivered inaccurate responses in seven out of 13 domains and inconsistent responses in seven out of 13 domains for drugs in all five classes of T2DM medication. Of ChatGPT’s responses to the 26 basic T2DM treatment questions, 7 (26%) were incorrect. In this cross-sectional study, we identified that it was common for ChatGPT to provide incorrect or inconsistent responses to enquiries related to the management of type 2 diabetes.

## Introduction

Large language models (LLMs) have fundamentally changed how patients and clinicians retrieve information.^1^ ChatGPT, the first widely available LLM-based chatbot, was released by OpenAI in 2022, and became one of the fastest-growing consumer applications in history. While LLM-based chatbots are now ubiquitous and accessible at no cost, evaluations of their accuracy for highly specialized medical knowledge are lacking.^2^ This is particularly true regarding medications for which knowledge is both specialized and dynamic.^3^ Our objective was to evaluate the accuracy and consistency of ChatGPT in answering questions related to the management of type 2 diabetes mellitus (T2DM). We chose T2DM for two reasons: First, T2DM affects over 11% of the global population.^4^ Second, over the past ten years, there have been multiple new breakthrough medications for the management of T2DM (e.g., sodium-glucose cotransporter-2 inhibitors and glucagon-like peptide-1 receptor agonists).^5^

## Methods

We conducted a cross-sectional study to evaluate the accuracy and consistency of the freely available version of ChatGPT as of June 2025 for (i) enquiries related to medications from the top five most common classes for T2DM medications and (ii) 26 basic questions related to the management of T2DM. The basic questions were drafted by our study team and are available in the Appendix. We selected ChatGPT because it is one of the most popular LLMs and among the top five in performance across widely used LLM benchmarks ^6,7,8^

Three team members (DC, TVB, PO) independently asked ChatGPT to formulate a table containing information for medications from each of the following classes: insulin, biguanides, dipeptidyl peptidase-4 (DPP4) inhibitors, sodium-glucose cotransporter-2 (SGLT2) inhibitors, and glucagon-like peptide-1 (GLP1) receptor agonists. The information requested covered the following 13 domains: mechanism of action, indication, route of administration, dosing regimen, absolute contraindications, common side effects, rare adverse effects, renal dosing, hepatic dosing, geriatric dosing, pregnancy considerations, black box warning, and patient counselling. We evaluated the consistency and accuracy of ChatGPT’s responses. Consistency was measured across all three responses in the same domain for a given medication. A response was labelled inconsistent if the response provided to one user differed from the response provided to at least one other user in the same domain for the same medication. Inaccuracy was measured by individual response. A response was labelled as inaccurate if the information provided by ChatGPT was incorrect based on the most recent FDA-approved drug label, in addition to review by an expert reviewer (MF).

The 26 basic questions related to the management of T2DM were categorized as correct or incorrect based on review by an expert reviewer (MF). We also reported the length of ChatGPT’s responses. We summarized all results using descriptive statistics. Research ethics board approval was not required because all of the data sources are publicly available.

## Results

ChatGPT delivered inconsistent responses in seven out of 13 domains for drugs in all five classes of T2DM medication. The most common domains for inconsistent responses (i.e., inconsistent in all five classes) were absolute contraindications and rare adverse effects. For example, for rare adverse effects of dapagliflozin (an SGLT2 inhibitor), the three responses provided were as follows:

Response to user 1: diabetic ketoacidosis.

Response to user 2: acute kidney injury, severe hypoglycemia.

Response to user 3: euglycemic DKA, Fournier’s gangrene.

ChatGPT also delivered inaccurate responses in seven out of 13 domains for drugs in all five classes of T2DM medication. The most common domains for inaccurate responses (i.e., inaccurate in all five classes) were absolute contraindications, common adverse effects, rare adverse effects, and pregnancy considerations. For example, for renal dosing considerations for metformin (a biguanide), the three responses provided were as follows:

Response to user 1: No adjustment required.

Response to user 2: Adjusted based on GFR.

Response to user 3: Reduce dose if eGFR 30-45; contraindicated <30.

Of ChatGPT’s responses to the 26 basic T2DM treatment questions, 7 (26%) were incorrect. The median word count of the responses was 212 words (interquartile range 167 to 304).

## Discussion

In this cross-sectional study, we identified that it was common for ChatGPT to provide incorrect or inconsistent responses to enquiries related to drugs from the top five most common classes of medications for the treatment of T2DM. We also identified that ChatGPT provided incorrect responses to one-quarter of the basic T2DM questions posed, and that the average length of response was approximately 200 words. For example, ChatGPT’s response to the question “Which diabetes medications offer cardiorenal protection?” contained a whopping 329 words. This was surprising, because this question—and many of the other questions posed—could be answered with a single sentence. Taken together, these results cast doubts on the suitability of ChatGPT for fielding enquiries related to medicine, and specifically to T2DM.

The high occurrence of inaccurate information is particularly concerning because patients and trainees may not be able to identify when a given response is inaccurate. ChatGPT and other widely available chatbots provide responses that are eloquent and delivered with confidence, regardless of the accuracy of the content. Our results suggest that chatbots being used for medical questions should ideally be trained on highly curated data that has been reviewed by medical experts.

Another concerning finding from our study was the high frequency of inconsistent responses provided to the same questions asked by different users. Because LLMs are probabilistic as opposed to deterministic, the inconsistency is to be expected. However, the end-user might not be aware of this, and such inconsistencies could have serious consequences if the information given is used for clinical decision-making.

The verbosity of ChatGPT’s responses to basic T2DM questions was expected. And while lengthy responses are not inherently concerning, often clinicians and trainees prefer relatively brief responses, wherein only the most relevant information is provided. Lengthy responses force the end user to sift through extraneous content. This can be particularly challenging for trainees, who are less experienced and may struggle to determine what information to prioritize when dissecting a lengthy response. Shorter responses markedly reduce this ambiguity by design.

Our results align with prior studies that have assessed the accuracy of ChatGPT for diabetes content. In a study by Barlas et al., it was found that ChatGPT’s responses were accurate on assessment of obesity in T2DM, but less accurate on the therapy-related questions, including questions on nutritional, medical, and surgical therapies for weight loss.^9^ In a study by Ying et al., researchers found that ChatGPT underperformed in correctness for more specialized information related to diabetes complications and treatment.^10^

Our study has a number of important limitations. First, we focused on enquiries related to type 2 diabetes, and thus, it is unknown how our results will generalize to other areas of medicine.

While we anticipate that the high occurrence of inaccurate and inconsistent information will be similar across other medical specialties, empirical data are needed to confirm. Second, we focused on ChatGPT exclusively, because of its widespread use; it is unknown how other similar LLM-based chatbots might perform. Third, the performance of LLMs continues to improve, and thus future versions of ChatGPT might have improved performance. Finally, we used the FDA label for each medication as the source of ground truth, but some labels contained inaccurate or outdated information. In such instances, we relied on expert opinion, which we acknowledge is subjective and imperfect.

## Conclusion

Our findings indicate that ChatGPT frequently delivers inconsistent or inaccurate responses to enquiries related to diabetes. These results underscore the importance of not relying on ChatGPT for clinical decision-making, and the need for LLMs that are fine-tuned using highly curated, up-to-date, accurate information, with expert oversight.

## Data Availability

All data sources are publicly available on ChatGPT.

## Appendix

### List of 26 questions

1. What is the first-line treatment for T2DM?
2. What is the A1C cut-off for diagnosing T2DM?
3. What are common side effects of metformin?
4. What are common side effects of DPP4i?
5. What are macrovascular complications of diabetes?
6. What are microvascular complications of diabetes?
7. Do both HbA1C and FPG need to be abnormal to diagnose T2DM?
8. Who should I screen for T2DM?
9. When should I start an SGLT2i in someone with T2DM?
10. When should I start a GLP1 in someone with T2DM?
11. How often should I screen my patient with T2DM for cardiac complications?
12. How often should I screen my patient with T2DM for peripheral neuropathy?
13. How often should I screen my patient with T2DM for kidney disease?
14. How often should I screen my patient with T2DM for retinopathy?
15. What should the blood pressure target be for a patient with T2DM?
16. When should I use a higher A1C target in my patient with T2DM?
17. How do I manage hypoglycemic episodes for my patients with diabetes?
18. How do I manage symptomatic hyperglycemia in my diabetic patient?
19. Which diabetes medications offer cardiorenal protection?
20. Do DPP4i cause weight gain?
21. Do sulfonylureas offer cardiorenal protection?
22. What is the Fasting Plasma Glucose and/or A1C for a patient considered at risk for diabetes?
23. What is the Fasting Plasma Glucose and/or A1C for a patient considered Prediabetic?
24. My patient has a diabetic foot. How do I know if it is osteomyelitis?
25. What is the gold standard to diagnose osteomyelitis in my diabetic patient
26. How common is diabetic retinopathy?

